# Multicenter benchmarking of short and long read wet lab protocols for clinical viral metagenomics

**DOI:** 10.1101/2024.01.14.24301284

**Authors:** F. Xavier Lopez-Labrador, Michael Huber, Igor A. Sidorov, Julianne R. Brown, Lize Cuypers, Lies Laenen, Bert Vanmechelen, Piet Maes, Nicole Fischer, Ian Pichler, Nathaniel Storey, Stefan Schmutz, Verena Kufner, Sander van Boheemen, Claudia E. Mulders, Adam Grundhoff, Patrick Blümke, Alexis Robitaille, Ondrej Cinek, Klára Hubáčková, Kees Mourik, Stefan A. Boers, Lea Stauber, Maud Salmona, Pierre Cappy, Alban Ramette, Alessandra Franze’, Jerome LeGoff, Eric C.J. Claas, Christophe Rodriguez, Jutte J.C. de Vries, European Society of Clinical Virology (ESCV) Network on Next-Generation Sequencing (ENNGS)

## Abstract

Metagenomics is gradually being implemented for diagnosing infectious diseases. However, in-depth protocol comparisons for viral detection have been limited to individual sets of experimental workflows and laboratories. In this study, we present a benchmark of metagenomics protocols used in clinical diagnostic laboratories initiated by the European Society for Clinical Virology (ESCV) Network on NGS (ENNGS).

A mock viral reference panel was designed to mimic low biomass clinical specimens. The panel was used to assess the performance of twelve metagenomic wet-lab protocols in use in the diagnostic laboratories of participating ENNGS member institutions. Both Illumina and Nanopore, shotgun and targeted capture probe protocols were included. Performance metrics sensitivity, specificity, and quantitative potential were assessed using a central bioinformatics pipeline.

Overall, viral pathogens with loads down to 10^4^ copies/ml (corresponding to C_T_ values of 31 in our assays) were detected by all the evaluated metagenomic wet-lab protocols. In contrast, lower abundant mixed viruses of C_T_ values of 35 and higher were detected only by a minority of the protocols. Considering the reference panel as the gold standard, optimal thresholds to define a positive result were determined per protocol, based on the horizontal genome coverage. Implementing these thresholds, sensitivity and specificity of the protocols ranged from 67 to 100% and 87 to 100%, respectively.

A variety of metagenomic protocols are currently in use in clinical diagnostic laboratories. Detection of low abundant viral pathogens and mixed infections remains a challenge, implying the need for standardization of metagenomic analysis for use in clinical settings.

## Introduction

Pathogen-agnostic metagenomic sequencing has emerged as a universal diagnostic method for infectious diseases^1^. This methodology allows for identification and genomic characterization of pathogens without a priori knowledge of a suspected pathogen. This approach is gradually changing the way physicians diagnose and manage infectious diseases^2^. In addition, pan-viral respiratory pathogen surveillance has been launched using metagenomic approaches^3^, enabling simultaneous tracking of all circulating viruses including potential novel ones, thus contributing to pandemic preparedness. The clinical utility of metagenomics in diagnosing idiopathic viral neurological syndromes has been reported in large prospective multi-center studies^4, 5^. However, implementation of metagenomics routinely in patient care has lagged behind^2^. Hurdles for widespread introduction in diagnostic settings include the complex and time-consuming workflows, the technical challenge of low biomass clinical samples such as cerebrospinal fluid, and the complicated interpretation of contaminating sequences. In addition, universal reference standards that mimic the high complexity of patient samples, and standardized approaches to demonstrate assay validation, are lacking^2^.

To date, reports on technical assessments of viral metagenomics protocols have been limited to individual sets of workflows and laboratories. Here, we present a benchmark study initiated by the European Society for Clinical Virology (ESCV) Network on NGS (ENNGS) including multiple metagenomic wet lab protocols used in clinical virology laboratories. A viral reference panel was designed to mimic low biomass clinical samples and used to assess the performance of twelve metagenomic wet lab protocols currently in use in diagnostic laboratories.

## Methods

### Construction of a reference panel

A viral metagenomic reference panel was designed to mimic low biomass clinical samples (e.g. respiratory swabs, cerebrospinal fluid) and their complexity, while reducing the number of environmental or cell culture-related sequences, to enable optimal sensitivity and specificity analyses. For this, twelve materials were prepared containing human cell free DNA (cfDNA, Twist pan-cancer reference standard set, 167 bp fragments with reduced methylation), spiked with synthetic viral sequences (both from Twist Bioscience, San Francisco, USA). These synthetic viral sequences covered >99.9% of the viral genomes of SARS-CoV-2 B.1.1429 Epsilon strain USA/CA-CZB-12943/2020 (EPI_ISL_672365), influenza A virus strain A/California/07/2009 (H1N1, NC_026438), measles strain Ichinose-B95a (NC_001498.1), and enterovirus D68 Fermon strain (NC_038308.1), in non-overlapping fragments of maximal 5 kb with 50 bp gaps for biosafety reasons according to the manufacturer’s policy. Viral sequences were mixed with several proportions of human cfDNA (90-99% of weight, up to 400 pg per 100 μl), corresponding with final proportions of 10-1% of viral nucleotides (down to 0.4 pg per 100 μl), based on the reported abundance in low biomass clinical samples^6, 7, 8, 9^. Concentrations of synthetic sequences were determined in triplicate by digital droplet PCR (BioRad QX200) and ranged from 10^4^ to 10^7^ copies/ml (cycle threshold, C_T_ values ranging from 24.4 to 31.1 in our assays^7, 10^). A virus negative cfDNA control was included.

In addition, two dilutions (1:100 and 1:1,000 in 0.1 Tris EDTA buffer) of ATCC Virome whole Virus Mix (MSA-2008™, ATCC, Manassas, USA) based on cultivated adenovirus (ADV) type F, cytomegalovirus (CMV), respiratory syncytial virus (RSV), influenza B virus, reovirus 3, and zika virus (C_T_ values from 27.8 to >40) were included in the panel. Viral loads of these dilutions were below the limit of quantification by the digital droplet PCR.

The panel was shipped to participants on dry ice, and receipt in good condition within 24 hours was confirmed by all sites. Nucleic acid (NA) extraction of the two ATCC Virome Virus Mix dilutions was performed locally (**Table 1**). Subsequently, all NA underwent library preparation according to local protocols (see below). An overview of the study design is shown in **Figure 1**.

**Figure 1.**
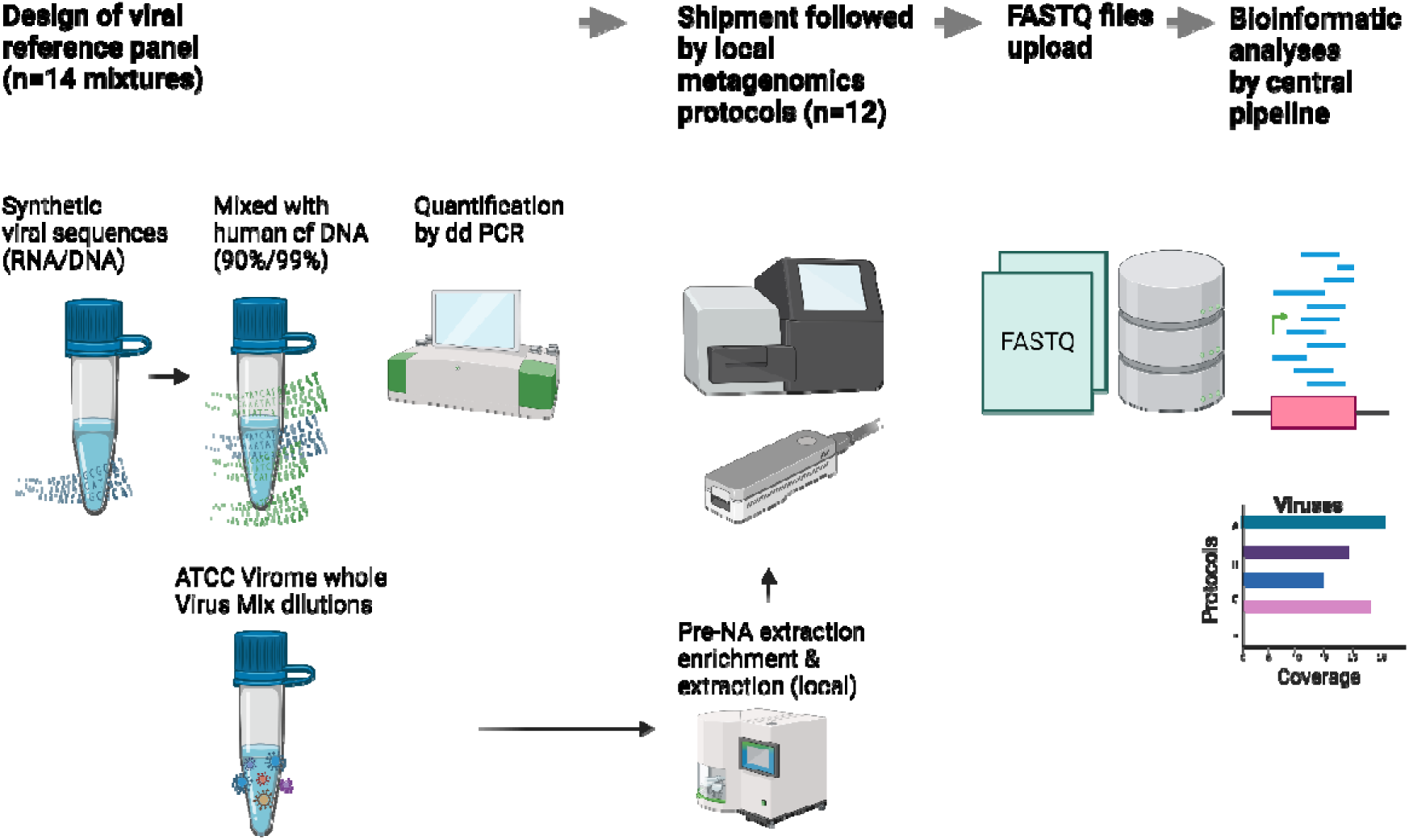
Overview of the study design. A mock virus reference panel was constructed of mixtures of synthetic virus sequences in human cell free DNA, and Virome whole Virus Mix dilutions. The panel was shipped to participating laboratories and sequenced by local metagenomic Illumina and Oxford Nanopore Technologies (ONT) protocols. Locally obtained raw FASTQ files were uploaded and analysed using a central bioinformatic pipeline. Created using Biorender.

**Table 1.** Protocol details of the metagenomics methods analyzed.

| Metagenomics protocol no. | 1 | 2 | 3 | 4 | 5 | 6 | 7 | 8 | 9 | 10 | 11 | 12 |
| --- | --- | --- | --- | --- | --- | --- | --- | --- | --- | --- | --- | --- |
| **Technology** | Illumina | Illumina | Illumina | Illumina | Illumina | Illumina | Illumina | Illumina | ONT | ONT | ONT | ONT |
| **Clinical use** | Experimental | Patient care | Patient care | Patient care | Patient care | Patient care | Experimental | Patient care | Experimental | Experimental | Experimental | Experimental |
| **In house/ commercial** | In house | In house | In house | In house | In house | In house | In house | Commercial | In house | In house | In house | In house |
| **Centrifugation filtration<sup>a</sup>** | No | No | No | No | 2000g 10 min, 0.45µm | No | 15,000g 3 min, no filtration | No | 17,000g 1 min, 0.8µm | 17,000g 1 min, 0.8µm | 5000g 5 min, 0.45µm | 2000g 10 min, 0.45µm |
| **Nucleic acid extraction<sup>a</sup>, Input/output volume** | ELiTe InGenius, ElitechGroup, 200 µl /100 µl | NA | 400 µl/ 60 µl | MagnaPure 96 DNA and Viral NA, 200 ul/100 ul | bioMérieux EMAG, 1000 µl/50 µl | easyMag NucliSENS/ specific B, 400 µl/60 µl | Qiagen Viral RNA kit, 140 µl/100 µl | MagnaPure 96 DNA and Viral NA, 200 ul/100 ul | QIAamp Viral RNA mini kit (Qiagen) | QIAamp Viral RNA mini kit (Qiagen) | Roche High Pure RNA kit, 100 µl/30 µl | bioMérieux EMAG, 1000 µl/50 µl |
| **Input volume for prep** | DNA: 24 µl<br>RNA: 24 ul | DNA: NA<br>RNA: NA | DNA: 26 ul<br>RNA: 10 ul | DNA: 50µl<br>RNA: 8 µl | DNA: 5 µl<br>RNA: 10 µl | DNA: 25µl<br>RNA : 11 µl | 12 ul (in 3 reactions) | 5 ul | 2.8 µl | 4 µl | 22 µl (in 2 reactions) | DNA: 5 µl<br>RNA: 10 µl |
| **rRNA depletion** | No | No | No (tissues only) | Yes | No | Yes | No | No | No | No | No | No |
| **Human DNA depletion** | No | No | No (tissues only) | No | No | No | No | No | No | No | No | No |
| **Library prep. kit DNA** | Illumina DNA prep (Nextera DNA flex), tagmentation | TruSeq DNA | NEBNext Ultra II DNA Prep | NEB Next Ultra II DNA | Nextera XT DNA Prep | TruSeq DNA | Combined DNA/RNA | Combined DNA/RNA: Twist Bioscience | Combined DNA/RNA: ONT Native Barcoding 96 V14 | Combined DNA/RNA: ONT Native Barcoding 96 V14 | Combined DNA/RNA: ONT PCR Barcoding Kit | PCR Barcoding Kit |
| **Library prep. kit RNA** | See DNA prep, incl. ds-cDNA synthesis | TruSeq Stranded Total RNA | KAPA RNA HyperPrep | SMARTer Stranded Total RNA-Seq | Nextera XT DNA Prep | TruSeq Stranded Total RNA | Nextera XT |  |  |  |  | ONT PCR Barcoding Kit |
| **Un/targeted** | Untargeted | Untargeted | Untargeted | Untargeted | Untargeted | Untargeted | Untargeted | Targeted: capture probes | Untargeted | Untargeted | Untargeted | Untargeted |
| Random amplification cycles | 12 | 12 | 8: DNA prep.<br>12: RNA prep. | 12 | 12 | 12 | 30 (SISPA) | 12 | 20 (WTA) | 30 (SISPA) | 30 | 45 |
| Internal controls spiked | - | - | RNA: phage MS2 | - | DNA: phage T1, RNA: phage MS2 | DNA: phage T1, RNA: phage MS2 | - | DNA: PhHV RNA: EAV | - | - | - | DNA: phage T1, RNA: phage MS2 |
| Total reads/sample (median), platform | 1.5M | 39M | 8M NextSeq 550 | 6.2M NextSeq500/2000 | 1.5M MiSeq | 5.6M | 1M NextSeq | 1.6M NovaSeq6000 | 1.8M P2 Solo | 3.8M P2 Solo | 0.1M | 0.6M Flongle, GridION |
| Volume (concentration) per library added to sequencing pool | 120-200 µl (8 pM) | NA | 0.9-13.5 µl | NextSeq 500: 40.5 µl (0.9 pM)<br>NextSeq 2000: 1.9 µl (400 pM) | 1.8 - 5 µl | 2 µl (4 nM) | NA | 19µl (30 nM) | 2.1 µl | 2.1 µl | 0.2-1.6 µl | 2.5 µl |
| References | Gradel et al<sup>20</sup><br>Bigi et al<sup>21</sup><br>Gradel et al<sup>22</sup> | Rodriguez et al<sup>23</sup> | Penner et al<sup>24</sup><br>Morfopoulou et al<sup>25</sup><br>Atkinson et al. 2023<sup>18</sup> | Alawi et al<sup>26</sup> | Kufner et al<sup>5</sup> | Narat et al<sup>27</sup><br>Vallet et al<sup>28</sup><br>Garzaro<sup>29</sup> and Aboudar et al<sup>30</sup> | Cinek et al<sup>31</sup> | Mourik et al<sup>19</sup><br>Carbo et al<sup>11</sup><br>Reyes et al<sup>32</sup><br>De Kleine et al<sup>33</sup> | Vanmechelen et al<sup>34</sup> | Wollants et al<sup>35</sup><br>Vanmechelen et al<sup>36</sup> | Van Boheemen et al<sup>6</sup> | Pichler et al<sup>37</sup> |

a; pre-nucleic acid extraction enrichment was applied to the ATCC Virome Mix dilutions containing whole viruses. WTA; whole transcriptome amplification, SISPA; sequence-independent, single-primer amplification. NA; not available

### Metagenomic protocols

In total, twelve metagenomic wet lab protocols were performed using the designed reference panel. The protocols were in use in the diagnostic laboratories of the participants. An overview of the protocol details and clinical use is shown in **Table 1**.

### Bioinformatic analysis

Raw FASTQ datasets obtained in the participating laboratories were uploaded at https://veb.lumc.nl/CliniMG (hosted by the department of Medical Microbiology at the LUMC, Leiden) and analyzed using a previously validated^11, 12, 13, 14^ central bioinformatics pipeline to exclude variation introduced based on differences in bioinformatic analyses. Non-inferiority of the central pipeline was ensured through comparison with target virus results as obtained by the corresponding local pipelines, using a criterium of 100% correspondence for qualitative detection of target viruses. Details on the local pipelines can be found in **Suppl. Table 1** and a previously published ENNGS benchmark study of pipelines ^14^.

After central quality pre-processing and removal of human reads by mapping them to the human reference genome GRCh38 (https://www.ncbi.nlm.nih.gov/assembly/GCF_000001405.26/ using Bowtie2^15^ version 2.3.4), datasets were analyzed using Genome Detective^16^ version 2.48 (accessed April – May 2023) as described previously^12^. Oxford Nanopore Technologies (ONT) datasets were subjected to QC using Genome Detective software. Genome Detective includes de novo assembly, and both nucleotide and amino acid-based classification in combination with a RefSeq / Swiss-Prot Uniref database^16^. Additional (off-target) viral classifications were confirmed by BLAST^17^.

### Performance metrics and statistical analyses

Both qualitative and quantitative performance of the protocols were analyzed based on horizontal genome coverage and sequence read counts. Sensitivity and specificity were calculated considering the reference panel as the gold standard; additional findings were considered false positives. Non-vertebrate viruses and endogenous retroviruses were excluded from analyses. Optimal thresholds to define a positive result were determined per protocol by varying the percentage of horizontal genome coverage, depicted in Receiver Operating Characteristic (ROC) curves. Target viruses up to PCR C_T_-values of 35 were included in the analyses. Read counts were normalized for total read counts (number of reads after QC per million, RPM), and for genome size for quantitative comparison with PCR C_T_-values using the formula: reads per kilobase per million (RPKM) = (number of reads mapped to the virus genome * 10^6^) / (total number of reads * length of the genome in kb)^12^. In case of separate RNA and DNA libraries, given the variability in size, total read counts of the corresponding separate libraries were used.

## Results

### Metagenomic wet lab protocols

The designed low biomass reference panel mimicking patient samples was sent to ten diagnostic laboratories. In total, twelve different viral metagenomic protocols were in use in the participating laboratories. Protocol details and clinical use are shown in **Table 1**. Eight Illumina and four ONT protocols were included. One of the methods targeted vertebrate viruses by probe hybridization, all other methods were untargeted. Most Illumina protocols were used for patient care, while all ONT protocols had an experimental status. Enrichment before nucleic acid extraction by filtration was used in all ONT protocols, but in only one of the Illumina protocols evaluated. Separate DNA and RNA libraries, as opposed to combined libraries, were prepared in 6/8 Illumina protocols in contrast to 1/4 ONT protocols. None of the protocols made use of depletion of human CpG methylated DNA during library preparation. Two protocols depleted ribosomal RNA. Random amplification of 20 cycles or more was used in all ONT protocols, and one Illumina protocol (see **Table 1**), including sequence-independent, single-primer amplification (SISPA). The median sequence read counts generated per sample ranged from 1 M to 39 M for Illumina protocols, and from 0.1 M to 3.8 M for ONT protocols.

### Detection of viral pathogens

Locally obtained FASTQ files were sent to the coordinating site for bioinformatic analyses. Performance of the metagenomic protocols was analyzed using a validated central pipeline that enabled processing of both short and long sequence reads. Overall, comparison of the results obtained by the central and local pipelines confirmed non-inferiority of the central pipeline in relation to the local pipelines as part of the overall local workflow for qualitative detection (**Suppl. Table 1**).

Qualitative detection of target viruses was first analyzed using the absolute, unnormalized data, representing the practical performance of the protocols with their corresponding platforms. A primary outcome parameter was selected that facilitated optimal comparison of short and long read protocols: the coverage percentage of the target virus genomes (% horizontal genome coverage, see **Table 2**). No thresholds for defining a positive result were used; that is, qualitative detection results by horizontal genome coverage and unnormalized read counts were therefore equivalent.

**Table 2.**
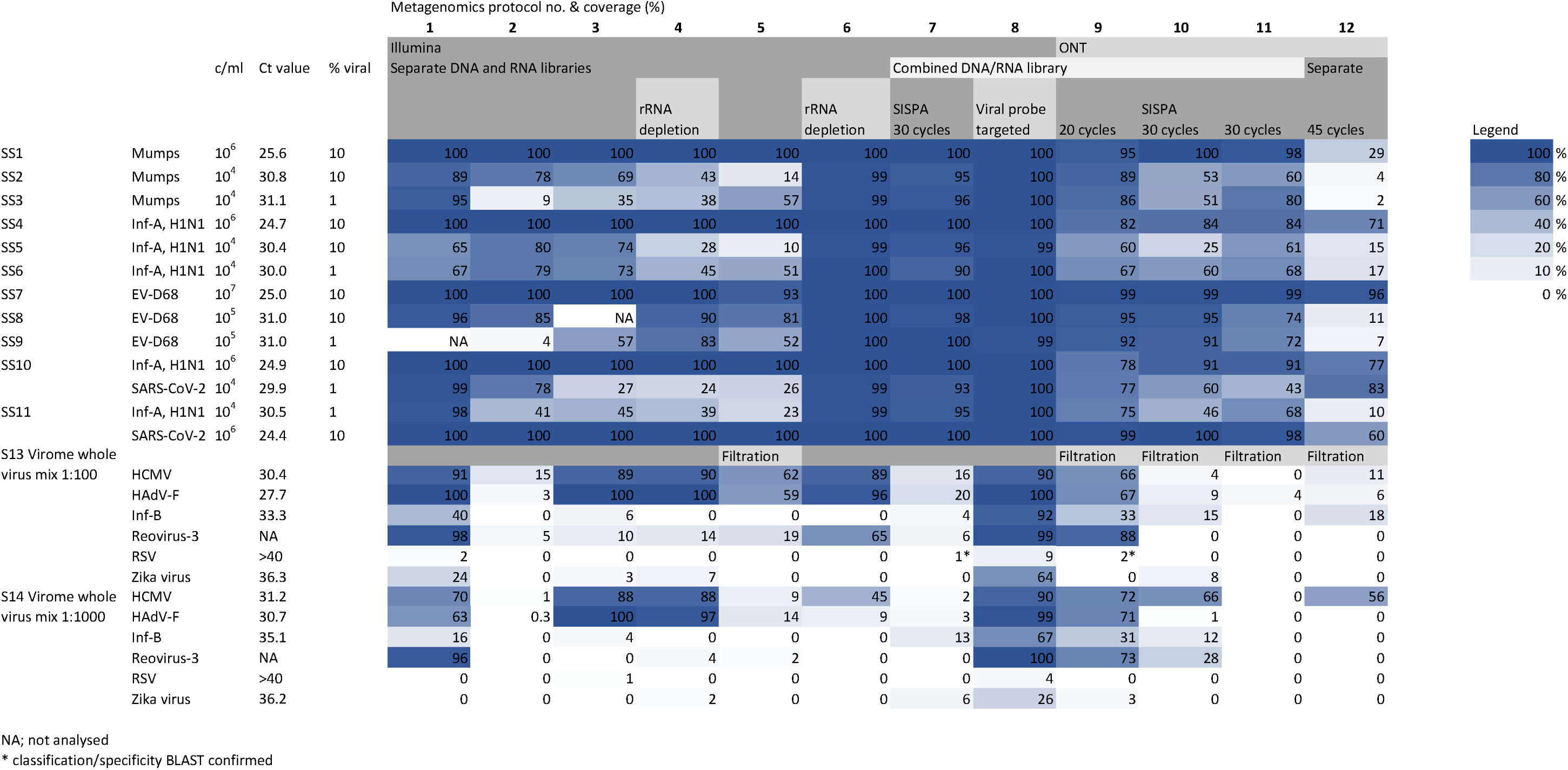
Detection of target viruses using <u>absolute</u>, unnormalized data, representing the practical performance of the protocols with their corresponding platforms. The <u>coverage percentage</u> of the target virus genomes is depicted, enabling comparison of short and long read protocols. No thresholds for defining a positive result were used.

Using the central pipeline, all protocols (12/12) resulted in 100% qualitative detection of the synthetic viral sequences spiked in up to 99% human background sequences (viral loads 10^4^-10^7^ copies/ml, C_T_ values 24-31). Coverages of the viral genomes were consistently 100% for viral loads of 10^6^ copies/ml or higher (C_T_ values of 24-26) when using the Illumina protocols, and ranged from 29% to 100% for the ONT protocols. Viral sequences with C_T_ values of 30-31 resulted in overall coverage of genomes of 95-100% for four of the Illumina protocols, coverage by the remaining Illumina and ONT protocols was lower.

Viruses present in the ATCC Virome whole Virus Mix up to C_T_ values of 31 were detected by all Illumina protocols and 3/4 ONT protocols. Viruses in the mix with C_T_ values of 33 to 35 were detected by 7 to 6/12 of the methods.

As secondary outcome measure, normalized read counts were compared to study the efficiency of the protocols with regard to sequencing target virus genomes in relation to overall sequences generated. Detection of target viruses based on normalized read counts (RPM) is shown in **Table 3**. Two of the four ONT protocols resulted in total read counts of median 1.0M and higher (**Table 1**). Detection based on RPM varied significantly among Illumina protocols and ONT protocols in a pattern distinct from detection based on genome coverage presented in **Table 2**. Highest RPM counts were obtained using the virus probe targeted protocol (#8, up to 0.9 M RPM), the Illumina protocol with separate DNA and RNA libraries and rRNA depletion (#6, up to 0.6 M RPM) and the ONT SISPA protocol (#10, up to 0.9 M RPM, though less consistent).

**Table 3.**
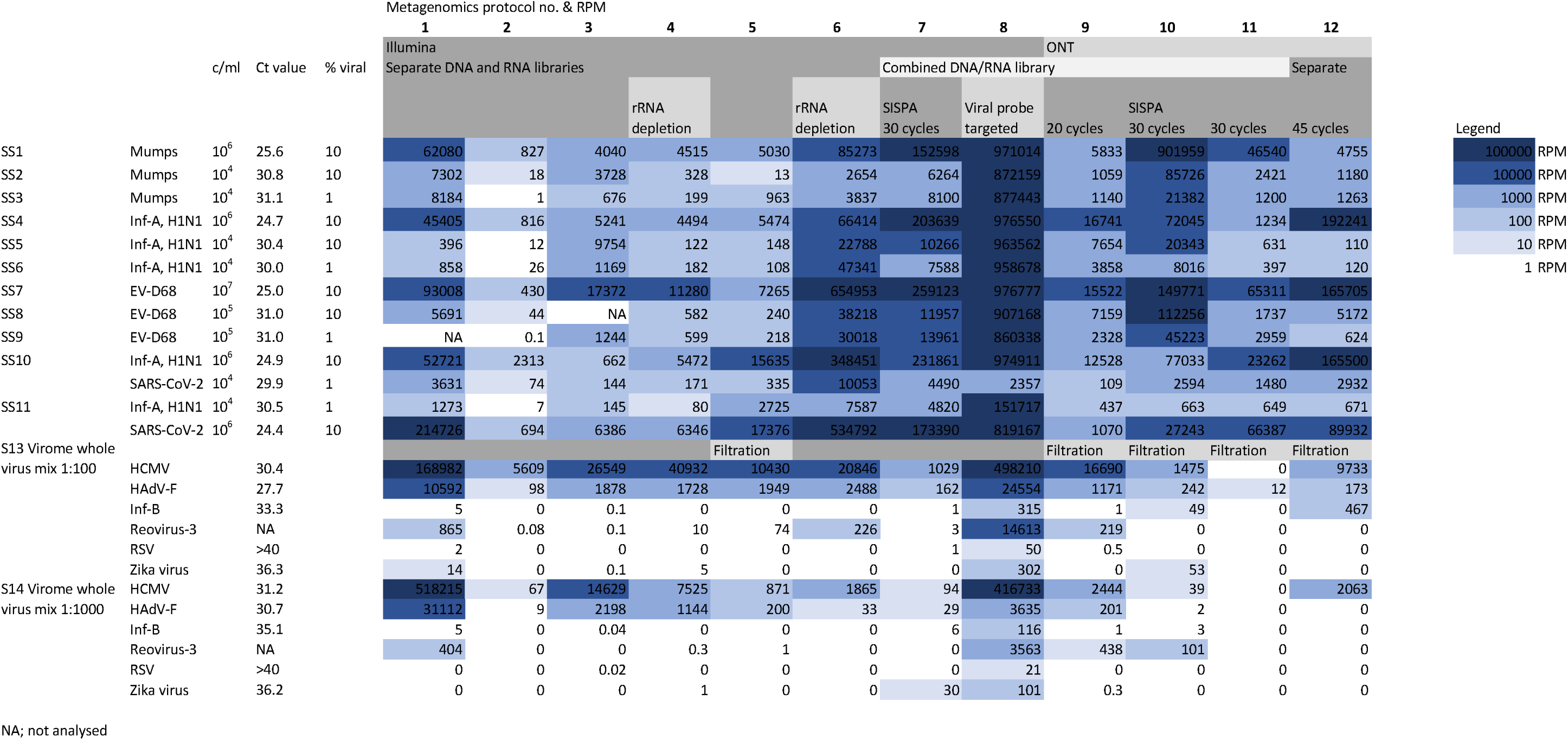
Quantitative results based on <u>normalized read counts (RPM)</u> for the target virus genomes in the panel, enabling comparison of efficiency of the protocols with regard to sequencing target virus genomes in relation to overall sequences generated. No thresholds for defining a positive result were used.

The correlation between sequence read counts (RPKM) and viral loads (copies/ml) ranged from 0.41 for the targeted protocol (#11) to 0.94 (#3, Pearson, **Figure 2)**. Illumina protocols generally had higher correlation coefficients than ONT protocols.

**Figure 2.**
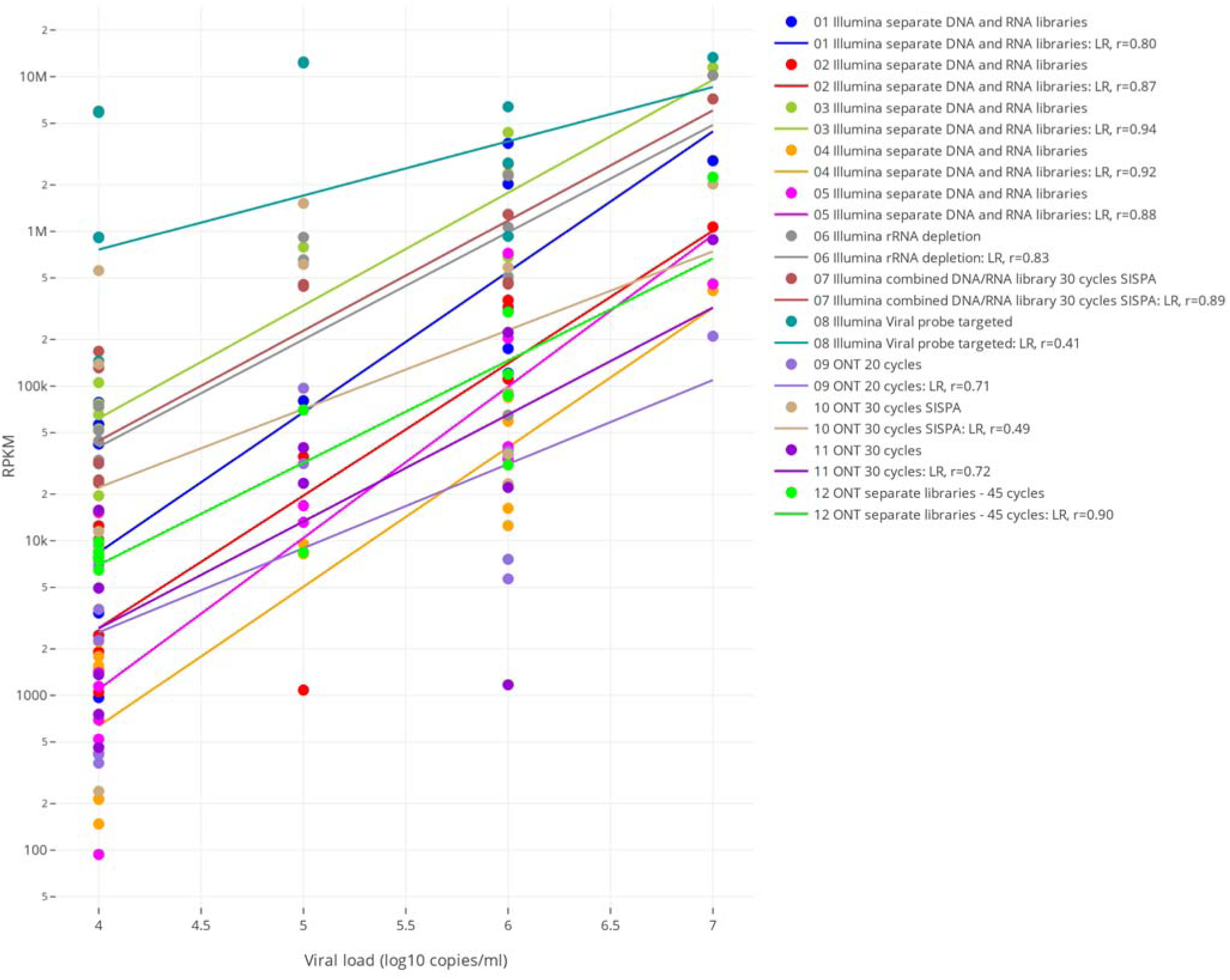
Correlation between normalized sequence read counts (per kilobase per million, RPKM) and viral loads (copies/ml) for the protocols. Legend: LR; linear regression, r; Pearson’s correlation coefficient, SISPA; sequence-independent single-primer amplification, ONT; Oxford Nanopore Technologies.

### Sensitivity, specificity and ROC curves

Sensitivity and specificity were calculated considering the reference panel as the gold standard; other detected viral sequences were considered false positives, and non-vertebrate viruses and endogenous retroviruses were excluded from analyses. Viral sequences detected by the central bioinformatics pipeline are listed per protocol in **Suppl. Table 2**, at species level and with their percentage of genomes covered. To enable comparison of the specificity, optimal thresholds to define a positive result were determined per protocol. Receiver Operating Characteristic (ROC) curves were generated by varying the threshold for the horizontal genome coverage in percentages, for all protocols (**Figure 3**). When considering the thresholds that resulted in optimal performance for each protocol, sensitivity varied from 67% to 100% (see legend of Figure 3). Specificity varied from 87-100%. Sensitivities and specificities of 95% and higher were obtained by four protocols: three Illumina (#1, #7, #8) and one ONT protocol (# 9).

**Figure 3.**
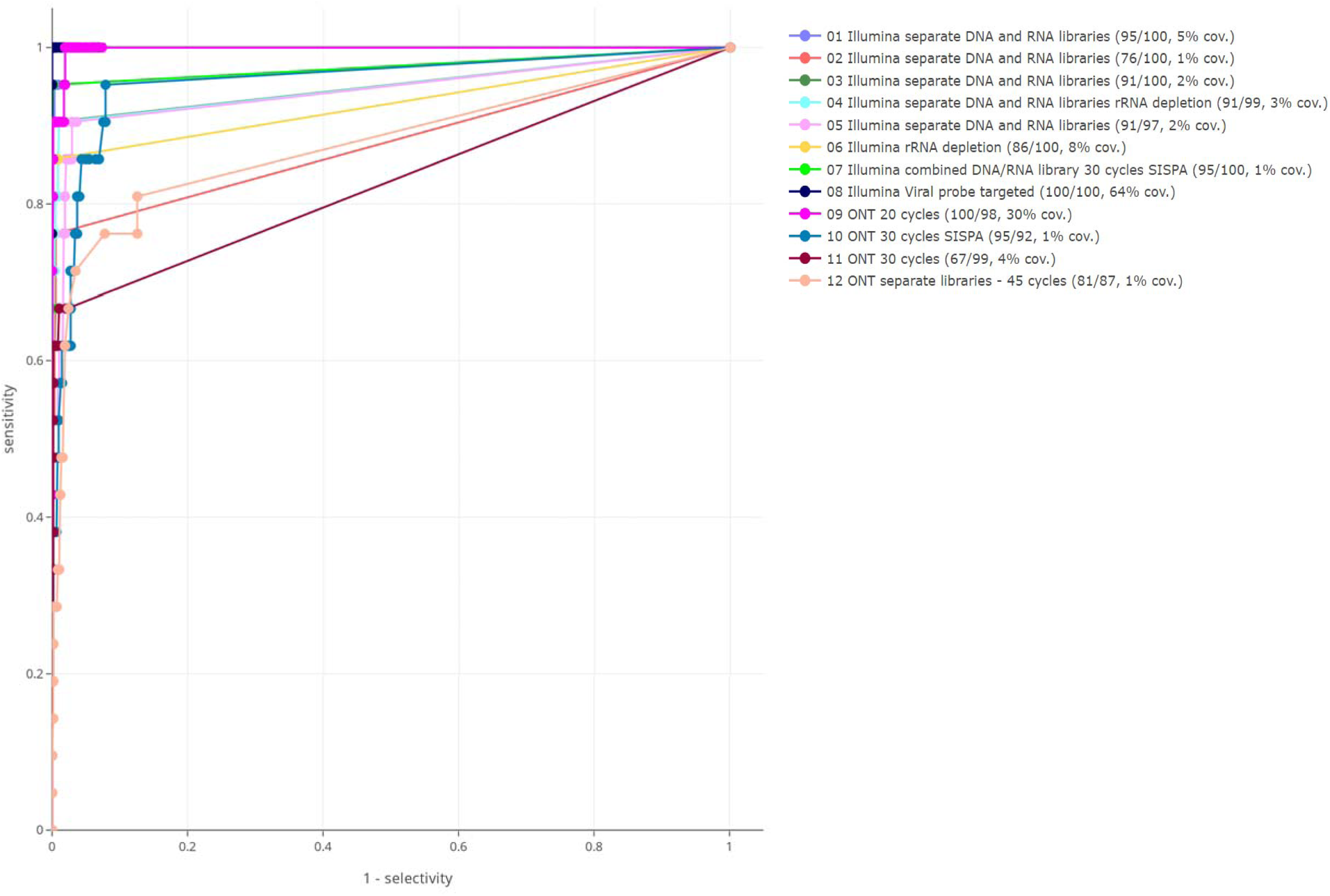
Receiver operating characteristic (ROC) curves based on varying threshold of percentage genome covered for defining a positive result for the twelve metagenomic protocols. Viruses up to PCR CT-values of 35 were included in the analyses. Legend: protocol name, sensitivity/specificity (%), and optimal threshold for horizontal genome coverage (% cov.) at the optimal ROC point.

A heatmap of the additional viral findings without taking into account these thresholds is shown in **Figure 4**, including the number of positive samples per species, and the maximum horizontal genome coverage in case of multiple hits per species. Additional findings could be classified as target virus contaminants (reference panel sequences), and off-target findings. The latter were mainly categorized as protocol specific (multiple samples with a single viral species hit within one protocol) but also shared off-target species (alphapolyomavirus, gammapapillomavirus) were detected by multiple protocols. On-target contaminants were slightly more often detected when using the most sensitive ONT protocols (#9 and #10). It cannot be excluded that the papillomaviruses and polyomaviruses were actually present in low concentration in the human background cfDNA originating from plasma.

**Figure 4.**
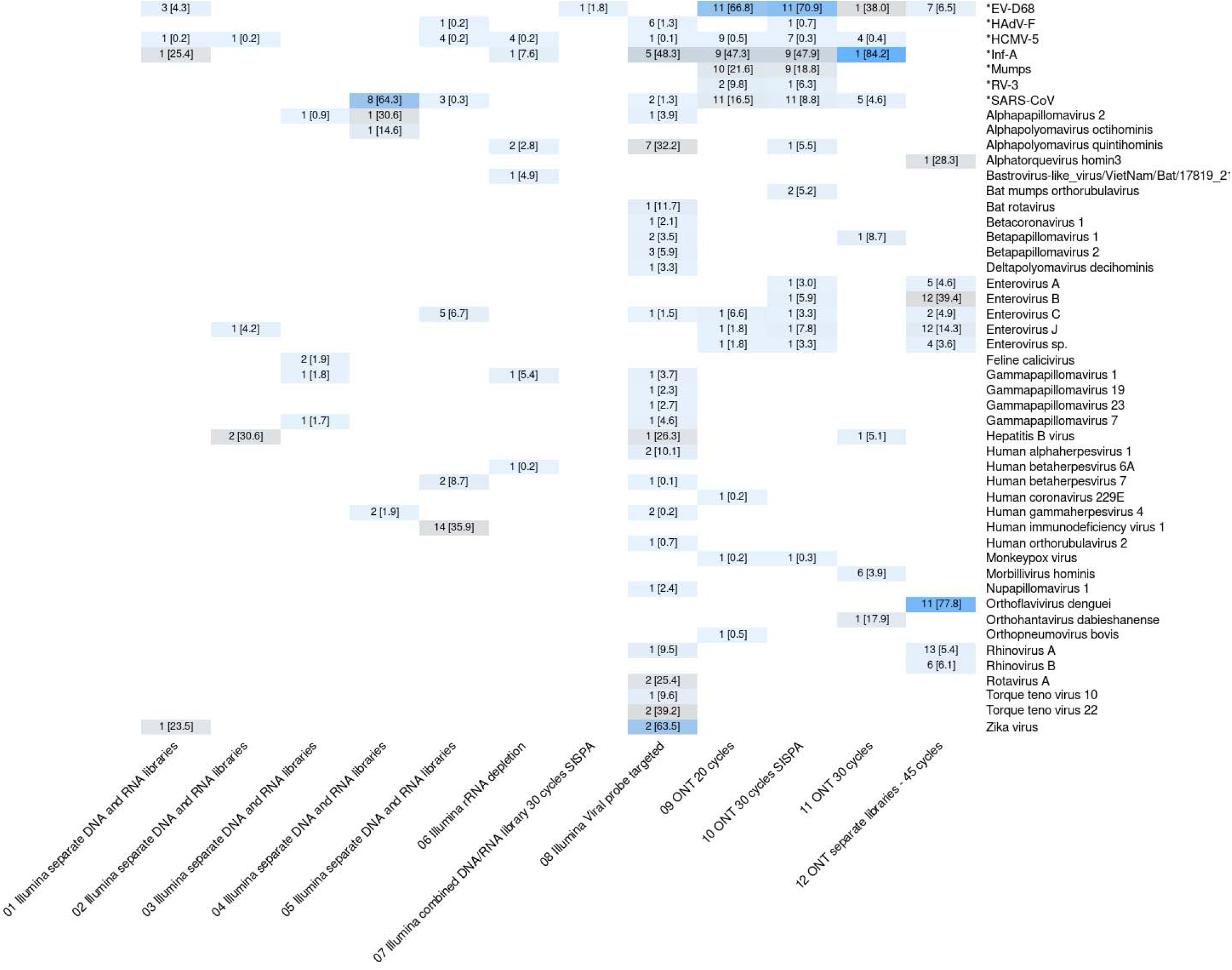
Additional viral species findings per protocol, without using thresholds for defining a positive result. Depicted are the number of pos. samples and [max. horizontal coverage] per finding (coloring for maximum coverage value range: ≤33%: pale blue, 33-66%: grey, ≥66%: blue). Pipeline classifications were confirmed by BLAST. Viruses marked with * are reference panel targets.

## Discussion

To our knowledge, this is the largest multicenter multinational in-depth cross-platform comparison of metagenomic wet lab protocols for viral pathogen detection reported to date. A low biomass reference panel containing 400 pg total nucleic acids per sample, below the common input recommended by manufacturers of routine library prep kits, was designed to mimic clinical sample types most commonly requested for metagenomics (cerebrospinal fluid) and most relevant for pandemic preparedness (respiratory swabs).

When introducing optimal per protocol thresholds, sensitivity of 90% and higher was accomplished using the majority of the protocols, and 10/12 protocols resulted in specificities of 95-100%. The protocols all reported correct qualitative results down to PCR C_T_ values of 31, suggesting that the performances of protocols were acceptable for clinical and surveillance settings using low biomass samples. However, the detection of contaminating sequences even in samples of a reference panel probably corresponded to ambient lab contamination, which remains a challenge for clinical metagenomic sequencing. The data presented illustrate efficient filtering out false positive findings when implementing protocol-specific thresholds for defining a positive result. Extraction methods may have a considerable impact on protocol performance. Lower viral load samples were included in the whole virus mixture without human background sequences, potentially affecting the efficiency of protocol steps and the clinical performance. Nonetheless, many of the undetected whole viruses were present at very low levels (CT values 33->40), close to or beyond the known limit of detection of some of the protocols^18, 19^.

A range of untargeted and targeted Illumina and ONT wet lab protocols were compared. Random amplification by SISPA (protocols #7, #10) resulted in higher normalized target read counts, however, this was not consistent for low viral load targets and did not result in improved horizontal genome coverage, indicating that amplification was not random over the entire viral genomes. The same phenomenon was seen for Illumina protocols that included either ribosomal RNA depletion or combined DNA and RNA libraries: the effect of higher coverages as compared to the other Illumina protocols was diminished when analyzing materials with low viral loads. It must be noted that the proportion of rRNA in this panel was low given a DNA only extraction that was used by Twist Bioscience to prepare the cfDNA, so the effect of an rRNA depletion step is less likely to be significant in this study setting. The hybridization targeted Illumina protocol (#8), however, consequently resulted in both higher genome coverage and normalized read counts.

This study has limitations. First, ideally, protocol comparisons are performed using clinical samples, but ring trials in general are limited by the available volume of clinical materials. We circumvented this limitation by using alternative materials mimicking clinical samples. Importantly, the reference panel enabled a reference for specificity analyses by providing a background that was relatively free from additional sequences. The panel provides a unique gold standard because it allows labeling of hundreds of viral species as true/false negative/positive, which would have been a tremendous effort to determine and quantitate by PCR in clinical samples. Second, by using nucleic acids as starting material to exclude potential effects of local nucleic acid extraction, pre-extraction enrichment steps could only be studied partially. Finally, the ssRNA and dsDNA reference pathogens included were aiming at pandemic preparedness and CSF syndromes, thus performance may not be entirely representative for other pathogens and all types of clinical materials.

To summarize, this collaborative work provided unique insight into the current state of implementation of viral metagenomic protocols for pathogen detection and the efficiency of a variety of wet lab protocol steps. In addition, we present a use case for a potential standardized bioinformatics validation approach using ROC curves with per protocol customized thresholds for calculation of specificity. This report aims to assist the implementation of viral metagenomics for pathogen detection in clinical diagnostic settings by providing insight into the efficacy of platforms and key protocol steps.

## Data Availability

All data produced in the present study are available upon reasonable request to the authors

## Acknowledgements

We thank David van der Meer (GenomeScan BV, Leiden) for performing ddPCRs.

